# COVID-19 Confirmed Case Incidence Age Shift to Young Persons Age 0-19 and 20-39 Years Over Time: Washington State March - April 2020

**DOI:** 10.1101/2020.05.21.20109389

**Authors:** Judith Malmgren, Boya Guo, Henry G. Kaplan

**Affiliations:** University of Washington, Dept. of Epidemiology, Seattle WA; HealthStat Consulting, Inc., Seattle WA; Swedish Cancer Institute, Seattle WA

## Abstract

**Background:** As the coronavirus (COVID-19) epidemic passes the peak infection rate in some states and counties a phased re-opening with changes of stay-at-home restrictions and social distancing recommendations may lead to an increase of nonessential work, social activities and gathering, especially among younger persons.

**Methods:** A longitudinal cohort analysis of Washington State Department of Health COVID-19 confirmed case age distribution March 1-April 19 2020 for proportional change over time using chi square tests for significance (N = 13,934).

**Results:** From March 1^st^ to April 19, 2020 age distribution shifted with a 10% decline in cases age 60 years and older and a 20% increase in age 0-19/20-39 years (chi-square = 223.10, p <.001). Number of cases over the eight-week analysis period were 0-19 years n = 515, 20-39 years n = 4078, 40-59 years n =4788, 60-79 years n = 3221, 80+ years n = 1332. New cases increased steadily among 0-19 and 20-39-year olds. After the peak (March 22, 2020), there was no decline among age 0-19 and a lesser decline among age 20-39 than older groups. As incidence declined in older age groups, the combined percentage of cases age 0-19 and 20-39 increased from 20% to 40% of total cases.

**Conclusions:** Increased COVID-19 infection among children and young adults is not without serious morbidity and mortality risk to them and others they may come in contact with, indicating a targeted approach for awareness and safety measures is advisable to reduce incidence among the supposedly less vulnerable but more mobile young population age 0-19 and 20-39 years.

## Introduction

The 2019 novel coronavirus (COVID-19) has quickly spread over the globe and positive cases continue to climb. The first reported case of 2019-nCoV infection was in Washington State on January 19, 2020 and on March 23^rd^, a statewide stay-at-home order was announced to remain in effect until May 4^th^ which has been extended to May 30, 2020.^1,2^ Change of stay-at-home restrictions and social distancing recommendations may lead to an increase of non-essential work, social activities and gathering, especially among younger persons. Using Washington State Department of Health (DOH) data, we analyzed incidence of COVID-19 cases by age for significant change in age distribution over time. As hospitalization rates declined and continued to decline but number of cases plateaued it became apparent age might be a factor to investigate to explain the changed presentation and severity of cases.

## Methods

Testing of symptomatic patients for COVID-19 in Washington State has been steadily increasing since the outbreak began in January 2020 with drive-through testing beginning in some communities in late March thus increasing testing availability. More than 25 labs in the state are now able to process COVID-19 testing. Total testing capacity statewide continues to change, which means each lab is the best source for current numbers on their own testing capacity. The results from all COVID-19 tests flow into the Washington Disease Reporting System (WDRS), an electronic disease surveillance system that allows public health staff in Washington state to receive, enter, manage, process, track and analyze disease-related data. Aggregate testing data is updated and published daily on the WA State DOH website with an update by age and county weekly.

We used the weekly updated COVID-19 positive confirmed case data from January 16^th^ to April 19^th^, Washington State Department of Health (DOH), available to the public on their website with updated data added for trend in age distribution 4/26-5/3/2020.^3^ The number of lab-confirmed COVID-19 cases from hospital, intensive care units, emergency departments and outpatient testing are reported by acute care hospitals in Washington daily. The data from DOH includes the number of cases, deaths and hospitalizations by week, county and age groups. Additional publicly available data on number of tests performed was also used in our study to address possible bias of changed testing by age over time. Our analysis used data updated May 3 2020 at 2 PM.

Laboratory-confirmed county-assigned cases by state assigned age groups 0-19, 20-39, 40-59, 60-79 and 80+ years in Washington State were enumerated and plotted over time. Statistical analyses were performed using Excel for percentage calculations and an online chi square calculator was used for two-sided significance testing with a .05 level of signficance.^4,5^ Weeks for analysis were restricted to March 1 to April 19 2020 when a sufficient number of cases (20 or more) had accrued to be statistically relevant and to accommodate the two-week lag time in case reporting and confirmation to WDRS. Age 0-19 was included even though there were only 4 cases in the first analysis week but case counts were greater than 20 in following weeks. Chi-square tests were run comparing the first two weeks of increased cases (3/1/2020, 3/8/2020), the peak week (3/22/2020) and the most recent weeks (4/12/2020, 4/19/2020). Number of weeks were restricted to 3 groups and time from 3/1/2020 to 4/19/2020 to avoid the diminishment of statistical validity by multiple comparisons.

## Results

Total number of positive cases in Washington State reported from January 16^th^ to April 19^th^ excluding positive cases with unknown age (n=10) were 14,220 with 13,934 confirmed cases from March 1 to April 19 [0-19 = 515 (4%), 20-39 = 4078 (29%); 40-59 = 4788 (34%); 60-79 = 3221 (23%); 80+ = 1332, (10%)]. The four counties with the highest number of cases by rank order were King (n=5955), Snohomish (n=2300), Pierce (n=1300), and Yakima (n=1119). As the epidemic progressed and the curve flattened, fewer older and more young persons tested positive for COVID-19 with the percentage of total cases among age 0-19 and 20-39 doubling from 20% to 40%. Figure 1. There was an increase in cases age 0-19 years over time from four cases week 3/1/2020 to 83 cases week 4/19/2020 and 131 week 5/3/2020. Incidence among age 60 and older declined 55% off peak but 2039-year-old cases only declined 36% off the peak week (3/22/2020) and thereafter have remained relatively constant. Table 1. The chi-square test statistic for the comparison of confirmed cases by age across the three discrete time periods of weeks 3/1 - 3/8, 3/22, and 4/12-4/19, was 223.10, p<.001. Table 1. Plotting percentage COVID-19 confirmed cases by age including the two most recent weeks [4/26/2020 (n=1554), 5/3/2020 (n=1210)] the age shift trend to age 0-19 and 20-39 was sustained over time to 50% of total cases. Table 1 and figure 2.

**Figure 1.**
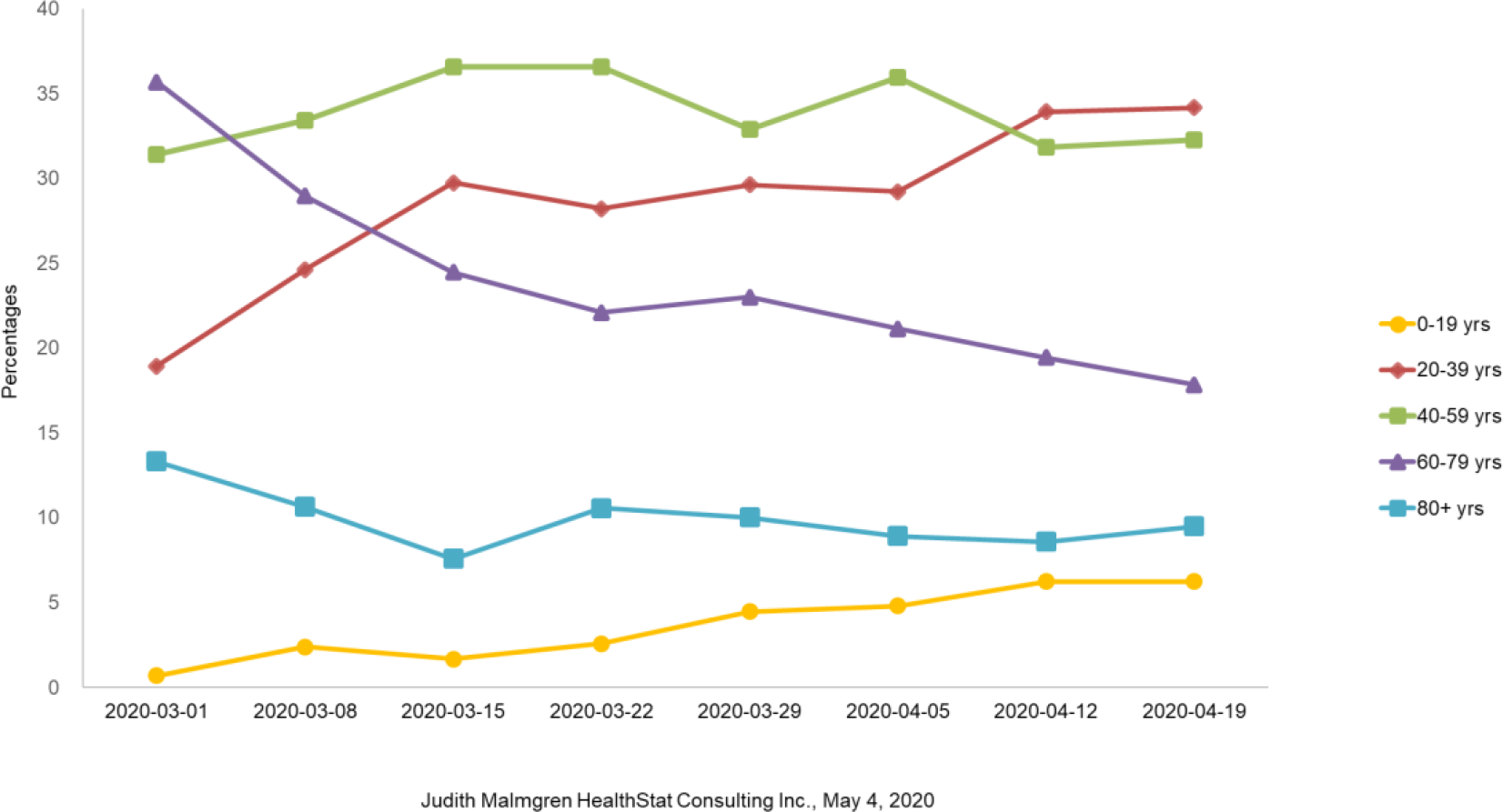
Washington State DOH Percentage of COVID-19 Confirmed Cases by Age: 3/1 - 4/19/2020, N=13,934

**Table 1.** WA State DOH COVID-19 confirmed cases by age: 3/1/2020-4/19/2020 (n=13934)

|  |  |  |  |  |  |  |  |  |
| --- | --- | --- | --- | --- | --- | --- | --- | --- |
|  | 0-19<br>N (%) | 20-39<br>N (%) | 40-59<br>N (%) | 60-79<br>N (%) | 80+<br>N (%) | Row<br>Total | Chi<br>Square | p-value |
| Week |  |  |  |  |  |  |  |  |
| 3/1* | 4<br>(.7%) | 111<br>(19%) | 184<br>(31%) | 209<br>(36%) | 78<br>(13%) | 586<br>(4%) | 223.10 | <.001 |
| 3/8* | 37<br>(2%) | 382<br>(25%) | 518<br>(33%) | 449<br>(29%) | 165<br>(11%) | 1551<br>(11%) |  |  |
| 3/15 | 38<br>(2%) | 680<br>(30%) | 836<br>(37%) | 559<br>(24%) | 178<br>(8%) | 2286<br>(16%) |  |  |
| 3/22* | 65<br>(3%) | 708<br>(28%) | 917<br>(37%) | 548<br>(22%) | 265<br>(10%) | 2509<br>(18%) |  |  |
| 3/29 | 101<br>(4%) | 669<br>(30%) | 743<br>(33%) | 554<br>(23%) | 226<br>(10%) | 2258<br>(16%) |  |  |
| 4/5 | 86<br>(5%) | 524<br>(29%) | 645<br>(36%) | 379<br>(21%) | 160<br>(9%) | 1794<br>(13%) |  |  |
| 4/12* | 101<br>(6%) | 550<br>(34%) | 516<br>(32%) | 315<br>(19%) | 139<br>(9%) | 1621<br>(12%) |  |  |
| 4/19* | 83<br>(6%) | 454<br>(34%) | 429<br>(32%) | 237<br>(18%) | 126<br>(9%) | 1329<br>(10%) |  |  |
| Column<br>total | 515<br>(4%) | 4078<br>(29%) | 4788<br>(34%) | 3221<br>(23%) | 1332<br>(10%) | 13934<br>(100%) |  |  |
| 4/26** | 131<br>(8%) | 542<br>(35%) | 531<br>(34%) | 219<br>(14%) | 131<br>(7%) | 1554 |  |  |
| 5/3** | 131<br>(11%) | 466<br>(39%) | 364<br>(30%) | 164<br>(14%) | 85<br>(7%) | 1210 |  |  |
| Column<br>Total** | 777<br>(4%) | 5086<br>(30%) | 5683<br>(34%) | 3604<br>(22%) | 1548<br>(9%) | 16698 |  |  |

\*\* Not included in the chi-square test analysis

**Figure 2.**
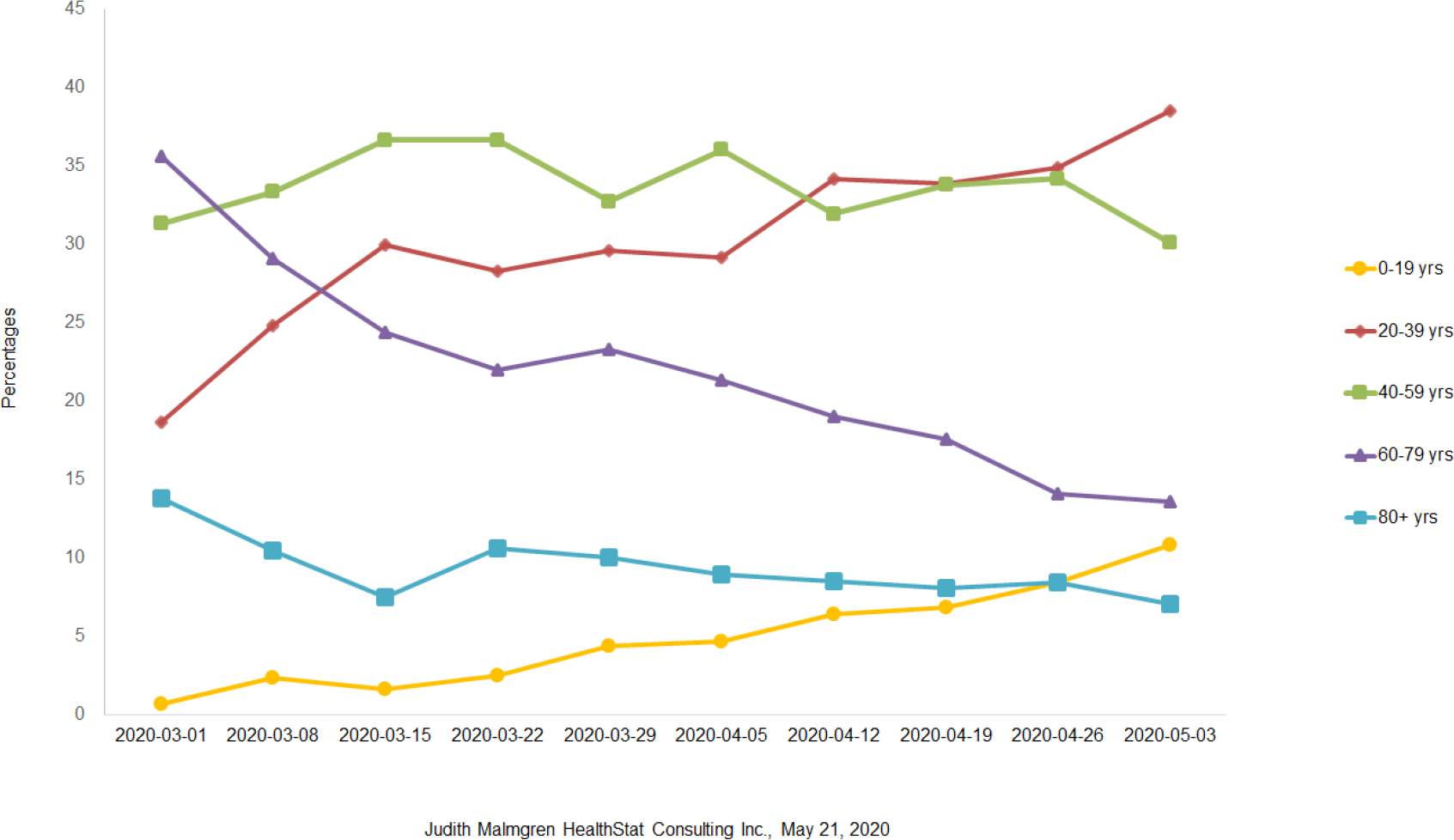
Washington State DOH Percentage of COVID-19 Confirmed Cases by Age: 3/1 – 5/3/2020, N=16,698

A total of 186,655 Covid-19 tests were run during the March 1 to April 19 2020 time period with 7.5% positive over the eight-week time period. Percent positive testing was variable over time and ranged from 12% (3/1/2020) to 5% (4/19/2020). Age distribution of persons tested changed by a small margin over time with an increase in testing among persons age 60 and older (+6%) and a decrease in testing among 0-19 and 20-39 year old (−3%) from the peak (3/22/2020) to the week of 4/19/2020, a statistically significant but small change [chi square = 263.87, p<.001]. Table 2.

**Table 2.** WA State DOH testing by age group: 3/1/2020-4/19/2020

| Table 2. WA State DOH testing by age group: 3/1/2020-4/19/2020 |  |  |  |  |  |  |  |  |
| --- | --- | --- | --- | --- | --- | --- | --- | --- |
| Week | 0-19<br>N (%) | 20-39<br>N (%) | 40-59<br>N (%) | 60-79<br>N (%) | 80+<br>N (%) | Total | Chi-square | P<br>value |
| 3/1 | 313<br>(16%) | 460<br>(24%) | 507<br>(26%) | 478<br>(25%) | 187<br>(10%) | 1945<br>(1%) | 263.87 | p<.001 |
| 3/8 | 2006<br>(13%) | 4911<br>(31%) | 4992<br>(31%) | 3162<br>(20%) | 863<br>(5%) | 15934<br>(9%) |  |  |
| 3/15 | 2231<br>(8%) | 9544<br>(35%) | 9041<br>(34%) | 5034<br>(19%) | 1098<br>(4%) | 26948<br>(15%) |  |  |
| 3/22* | 1832<br>(6%) | 10257<br>(36%) | 10000<br>(35%) | 5587<br>(19%) | 1177<br>(4%) | 28853<br>(16%) |  |  |
| 3/29 | 1825<br>(7%) | 9851<br>(35%) | 9611<br>(34%) | 5445<br>(19%) | 1331<br>(5%) | 28063<br>(15%) |  |  |
| 4/5 | 1681<br>(6%) | 9041<br>(34%) | 9235<br>(34%) | 5658<br>(21%) | 1273<br>(5%) | 26888<br>(15%) |  |  |
| 4/12 | 1612<br>(7%) | 7945<br>(32%) | 7807<br>(32%) | 5550<br>(22%) | 1753<br>(7%) | 24667<br>(13%) |  |  |
| 4/19* | 2094<br>(7%) | 9711<br>(31%) | 10572<br>(34%) | 7238<br>(23%) | 1732<br>(6%) | 31347<br>(17%) |  |  |
| Total | 13594<br>(7%) | 61720<br>(33%) | 61765<br>(34%) | 38152<br>(21%) | 9414<br>(5%) | 184645 |  |  |

From the COVID-19 peak week (3/22/2020) to week 4/19/2020, COVID-19 hospitalization rate declined 49% for age 40 and older cases with a 54% decline in COVID-19 incidence. The hospitalization rate among age 20-39-year-olds declined 35% with 36% decline in confirmed positive cases from the peak.^3^

## Discussion

Although morbidity and mortality from COVID-19 infection are highest among the infected population 60 years and older and those with underlying conditions, the percentage of 0-19 and 20-39-year-old cases in Washington State increased over time with no decline in cases age 0-19-year-old cases and a lesser decline among 20-39-year-old than observed in older age groups. Our results indicate a persistent high percentage of current infections in Washington State is among the age 0-19 and 20-39 population who may also be at highest risk of contracting and spreading the virus but not at high risk of hospitalization or mortality. During this time period, COVID-19 related hospitalization rates declined without an equivalent rate of decline among confirmed COVID-19 cases. The disproportionate rate of case hospitalization and confirmed case decline between older and younger age groups is concerning. The shift from from older to younger population COVID-19 infection may mask a true decline in cases and the need for future health care capacity if the currently infected portion of the population is younger, less likely to report symptoms, and at less risk of a severe life-threatening disease requiring hospitalization. In Seattle Washington 40% of the population is age 20-39 years old compared to 28% for the state and is located in the most populous county (King County 2019 population 2,252,782).^6^ From our observations persons age 20-39 years old may be a specific group to target for infection control behavior modifications and directives as they are more often employed in work sectors with high levels of public contact.

As initial public warnings targeted the population age 60 and older and those with underlying conditions, a misconception may be at large that only persons age 60 and older are at risk for contracting COVID-19. Children have been thought to be at low risk for COVID-19 and if infected to have a mild clinical course with few or no symptoms. A health alert has been issued by the CDC for Pediatric Multisystem Inflammatory System (MIS-C) linked to COVID-19 indicating in some cases a low risk assessment may not be accurate.^7^ The perception that age 0-19 and 20-39 age groups are not at high risk for morbidity and mortality from COVID-19 and thus might be free to interact with others discounts the possibility of asymptomatic transmission and thereby endangers older adults and those with comorbid conditions they come in contact with. Younger persons may be more likely to participate in work and social activities that involve prolonged and intensive contact with others. Findings from a recent survey by Canning et al, found persons under age 50 had twice the predicted number of close contacts than older people.^8^

### Limitations

While testing overall has increased during the period evaluated, the percentage of younger people being tested has decreased slightly compared to persons age 60 and older. Absent information to link family and household level data with parent’s occupation to identify the source of exposure in the 0-19-year-old age group, one could assume the infections are driven by contact with family members working outside of the home as Washington State public schools have been closed since March 11, 2020.

The Washington State stay-at-home policy is temporally associated with the observed case decline after March 22, 2020.^9^ Specific advisories to children and teenagers age 0-19 years and 20-39-year-old adults to increase awareness of COVID-19 transmission and infection among younger age groups may be advisable to reduce overall COVID-19 incidence levels and enhance movement towards levels that will allow phased reopening of the state and counties.^10^

Our findings indicate a justifiable concern regarding the phased reopening plan for Washington State in late May in light of the shift in COVID-19 incidence from older to younger age with the majority of current cases now in the combined 0-19/20-39-year-old age group. This specifically affects counties with a high percentage population age 20-39 years. Continued tracking of age trends are warranted and will help target which activities can be opened and when that might occur. Going forward, younger persons with possible greater communicable capacity especially in King County could be included in the priority for identifying, controlling and stopping the spread of COVID-19. As Washington State is the first in the United States with COVID-19 experience and the longest outbreak timeline, our experience indicates analysis of incidence by age and county are useful tools for measuring outbreak progression.

## Data Availability

The data used was publicly available deidentified data downloaded from the Washington State Department of Health COVID-19 Dashboard tab #2 for confirmed cases and hospitalizations
https://www.doh.wa.gov/Emergencies/NovelCoronavirusOutbreak2020COVID19/DataDashboard

## Acknowledgements

The authors wish to express their heartfelt thanks and gratitude for the invaluable assistance of Michelle Holshue, RN, MPH of the Washington State Department of Health and the CDC Epidemiologic Intelligence Service. The authors also wish to thank the Public Health staff in every county in Washington State and the Washington State Department of Health for their hard work collecting the data and their contribution to the Washington Disease Reporting System.

